# Estimating the overall effect of a mass drug administration strategy for malaria in Los Chiles, Costa Rica: One-year target trial emulation with synthetic controls

**DOI:** 10.1101/2025.08.01.25332751

**Authors:** Santiago Esteban, Adriana Ornelas, Adriana Torres Moreno, Rodrigo Marin Rodriguez, Melissa Ramirez Rojas, Isaac Vargas Roldan, Hazel Herra Bogantes, Jennyffer Gonzalez Luna, Yadel Centeno Ureña, Leopoldo Villegas, Diego Rios-Zertuche

**Affiliations:** Regional Malaria Elimination Initiative, Health, Nutrition and Population Division, Inter-American Development Bank, Buenos Aires, Argentina; Regional Malaria Elimination Initiative, Health, Nutrition and Population Division, Inter-American Development Bank, Mexico City, Mexico; Health Surveillance Direction, Costa Rica’s Ministry of Health, San José, Costa Rica; Vector Control Program, Costa Rica’s Ministry of Health, San José, Costa Rica; Huetar Norte Health Region, Costa Rica’s Ministry of Health, San José, Costa Rica; Los Chiles Health Area, Costa Rica’s Ministry of Health, Los Chiles, Alajuela, Costa Rica; Regional Malaria Elimination Initiative, Health, Nutrition and Population Division, Inter-American Development Bank, San José, Costa Rica; Regional Malaria Elimination Initiative, Health, Nutrition and Population Division, Inter-American Development Bank, Washington DC, USA; Independent Senior International Development Adviser, Silver Spring, Maryland, USA

## Abstract

**Background:** Mass drug administration (MDA) is a WHO-recommended strategy for the elimination of *Plasmodium falciparum* malaria in low-transmission settings close to elimination. After achieving zero autochthonous cases between 2013 and 2015, Costa Rica experienced a resurgence, primarily of *P. falciparum*, prompting an MDA intervention in 2023 in the Los Chiles focus. We evaluate its impact on the incidence of autochthonous malaria.

**Methods:** An MDA with two cycles of chloroquine (25 mg/kg per cycle, spaced seven weeks apart) was implemented between April—June 2023 in three localities (Medio Queso, Coquital, and San Gerardo) within the Los Chiles focus. We use a cluster-target trial emulation to estimate the overall average treatment effect on the treated (ATT) under an intention-to-treat approach. We use generalized synthetic control methods (GSCM) to construct a counterfactual for the intervened localities, using malaria surveillance data from January 2021 to April 2024. Our primary outcome was the autochthonous malaria case count at 1, 3, 6, and 12 months post-intervention.

**Results:** We found that the MDA achieved 93.3% coverage across the two cycles (4,316/4,624 people received at least one dose). The first cycle had 77.3% coverage and the second 51.2%, achieving 68.1% adherence (full treatment in at least one cycle) and 20.4% in both cycles. Following the intervention, autochthonous malaria cases in the treated localities decreased to zero in the first month and remained at zero throughout the 12-month follow-up period. Overall, we estimated that in the absence of MDA, cases would have persisted (approximately 3-4 cases/month). The strategy was associated with a significant reduction in cases, with an overall post-intervention Rate Ratio (RR) of 0.23 (95% CI: 0.13, 0.49) and an average reduction of 3.72 cases per period (95% CI: −7.2, −0.94) compared to the synthetic control.

**Conclusion:** We found that the chloroquine-based MDA strategy implemented in the Los Chiles focus was highly effective in interrupting autochthonous malaria transmission in the short and medium term. Our results suggest that MDA, when well-planned and implemented in a low transmission setting with a strengthened health system, can be a powerful tool to accelerate malaria elimination.

## Background

Mass drug administration (MDA) has high potential to accelerate elimination efforts of *Plasmodium falciparum* and *Plasmodium vivax*, especially in areas with low transmission [1]. MDA consists of administering the full antimalarial treatment course, targeted to the specific parasite, to all individuals within a defined population or geographic area, except for those for whom treatment is contraindicated, over the same time period and at repeated intervals [2]. Recently, the World Health Organization (WHO) recommended MDA for the elimination of *P. falciparum* in special situations, including the interruption of transmission in areas approaching elimination [2]. Its successful implementation depends on specific conditions: a health system with adequate surveillance, access to effective diagnosis and treatment, and robust vector control [1–5]. Furthermore, the risk for reestablishment of transmission must be minimal. Operationally, MDA requires achieving high population coverage (greater than 80%), often through repeated rounds, ensuring adherence to the full treatment course through strong community participation and engagement, and preferably being administered during the low transmission season [1–5].

Few studies have documented the long-term effects of MDA on interrupting malaria transmission. [6]. Furthermore, most MDA experiences have been documented in African countries, and only a few examples of MDA in Latin America and the Caribbean [7], especially under low transmission conditions. In Costa Rica, an MDA campaign was conducted in the Boca Arenal community, located in the Huetar Norte region, in January[8]. At that time, unlike current trends, the predominant malaria parasite was P. vivax. The campaign was conducted in a single round, during which participants received three doses of chloroquine (a total of 1,500 mg) and seven daily doses of primaquine (30 mg per day) for radical cure [8]. Among the eligible population, 92.1% received at least one dose, and 47.4% completed the full treatment regimen [8]. The MDA temporarily reduced transmission for approximately three months; however, transmission was later reestablished due to imported cases [8].

Several literature reviews investigated the effectiveness of MDA in interrupting malaria transmission [1,4,6,9–11]. A recent Cochrane review found that in low- and very low-transmission settings, MDA potentially reduces *P. falciparum* parasitemia and prevalence within three months [6]. Yet, the authors conclude that there is insufficient evidence that its effects are sustained beyond three months [6]. Another review, including gray literature, unpublished reports, and interviews with implementers, concluded that MDA can interrupt malaria transmission in areas of low endemicity, especially in relatively isolated areas with small populations and limited population mobility [1]. However, they also conclude that there are experiences where transmission was interrupted in more complex scenarios [1].

Central America is considered one of the regions with the greatest potential to eliminate *P. falciparum* and a high potential to eliminate *P. vivax* [12]. Costa Rica could be the next country to eliminate indigenous transmission of malaria. Between 2013 and 2015, Costa Rica achieved zero autochthonous cases [13], meaning cases that occurred due to transmission dynamics within the country itself and were not related to imported cases from other countries. However, transmission continued in 2016, resulting in 543 autochthonous cases by 2023 [13]. During this same year, 92% of autochthonous transmission cases were due to *P. falciparum,* and the remaining 8% due to *P. vivax*. Most cases, 86%, occurred in the Huetar Norte and Huetar Caribe regions [14], in the north and northeast of the country near the border with Nicaragua. The increased incidence of P. falciparum cases, localized in certain communities in these regions, motivated the Costa Rican Ministry of Health authorities to implement a new MDA intervention in April 2023.

### MDA Strategy in the Los Chiles Focus, Alajuela

The concentration of malaria cases in Los Chiles, within a relatively small area of the Huetar Norte Region (see *Fig 1*), provided the rationale to implement MDA. The Huetar Norte Region includes the cantons of San Carlos, Guatuso, Los Chiles and Upala in the Province of Alajuela, the canton of Sarapiquí in the Province of Heredia, the districts of Peñas Blancas in the canton of San Ramón, Río Cuarto in the canton of Río Cuarto and Sarapiquí in the canton of Alajuela. Epidemiological data from January 2021 to March 2023 show that the Los Chiles focus accounted for 80.4% of the region’s autochthonous malaria cases. Despite the intensity of transmission, the focus is classified as a low transmission setting, in line with the recommendations for MDA implementation [2].

**Fig 1.**
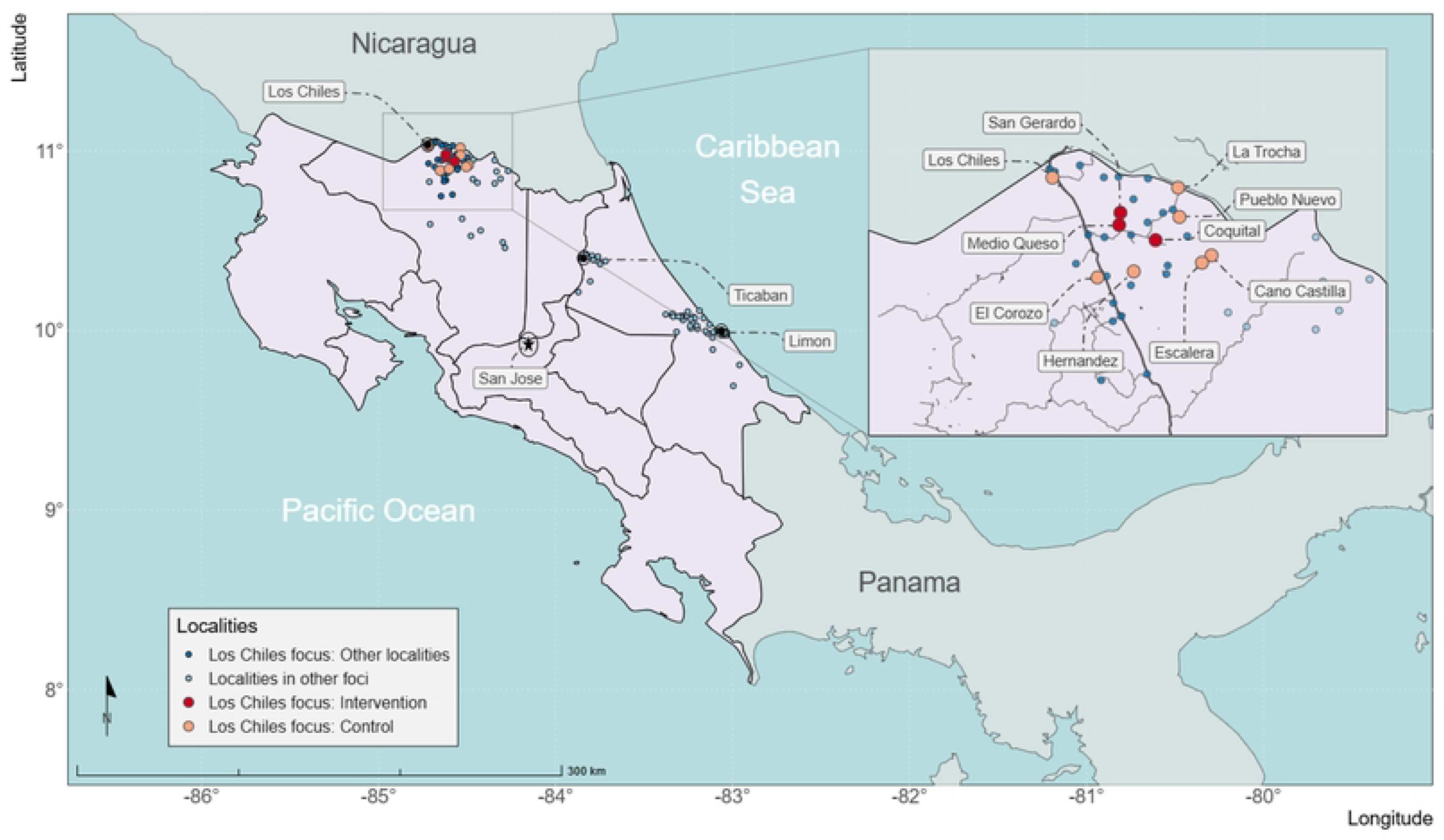
Map of intervention and control localities

In 2023, the Los Chiles focus area comprised 19 localities within the Los Chiles district, three in the district of El Amparo, and one in San Jorge. Together, these three districts represent 8.6% of the total population in the Huetar Norte region. Autochthonous malaria cases followed an increasing trend from 2021 to 2023, reaching a peak of 63 cases in October 2021 (see Fig 2). A sharp decrease in cases began in April 2023, coinciding with the start of the intervention.

**Fig 2.**
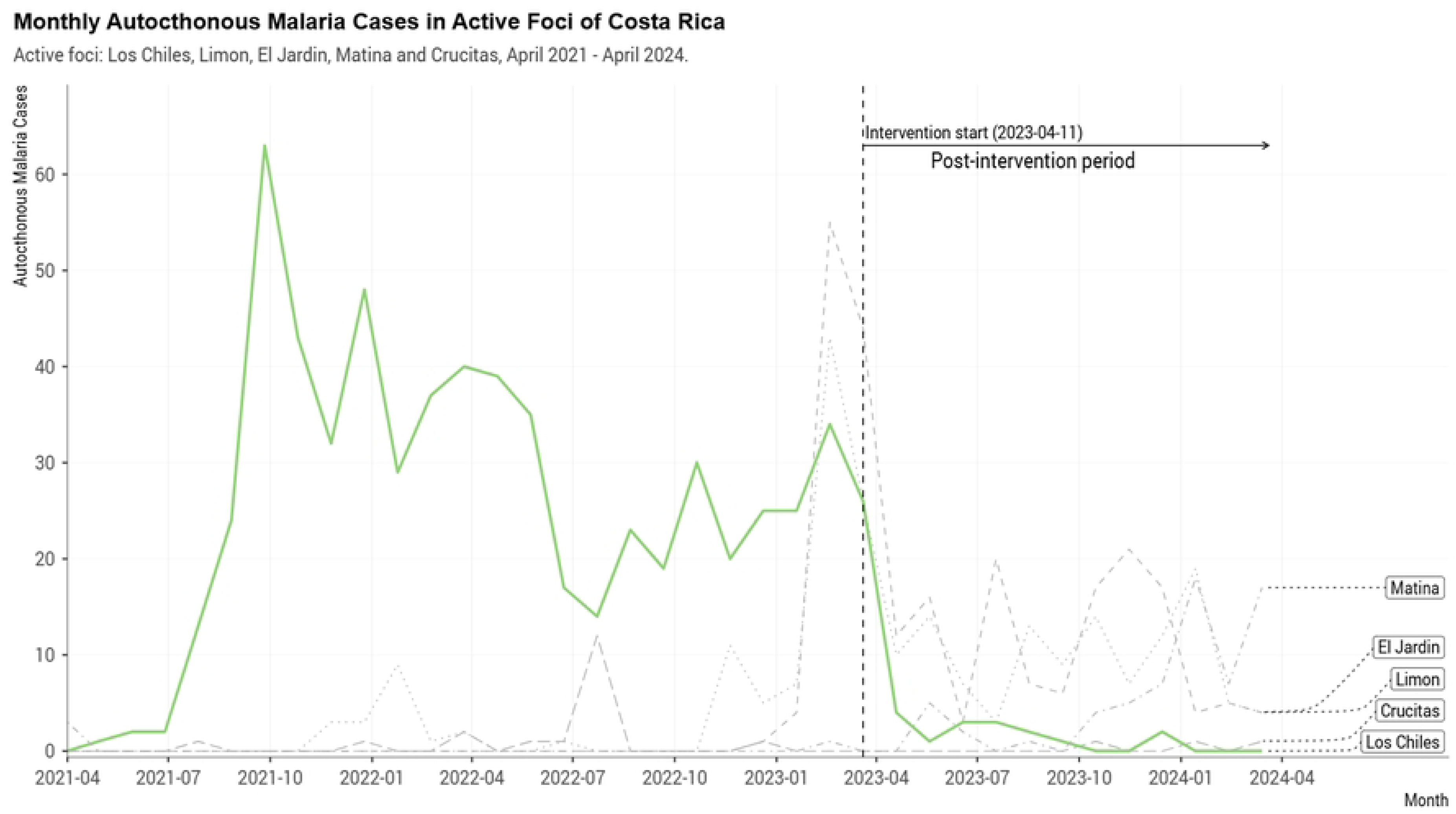
Autochthonous malaria cases reported in the foci of Los Chiles, Crucitas, El Jardín, Limón, and Matina in Costa Rica, April 2021–April 2024.

Between April 2021 and March 2023, *P. falciparum* was responsible for over 90% of malaria cases, significantly outnumbering *P. vivax*. Since 2021, the localities of Medio Queso, Coquital, and San Gerardo have reported the highest number of cases within the focus (see Fig 3).

**Fig 3.**
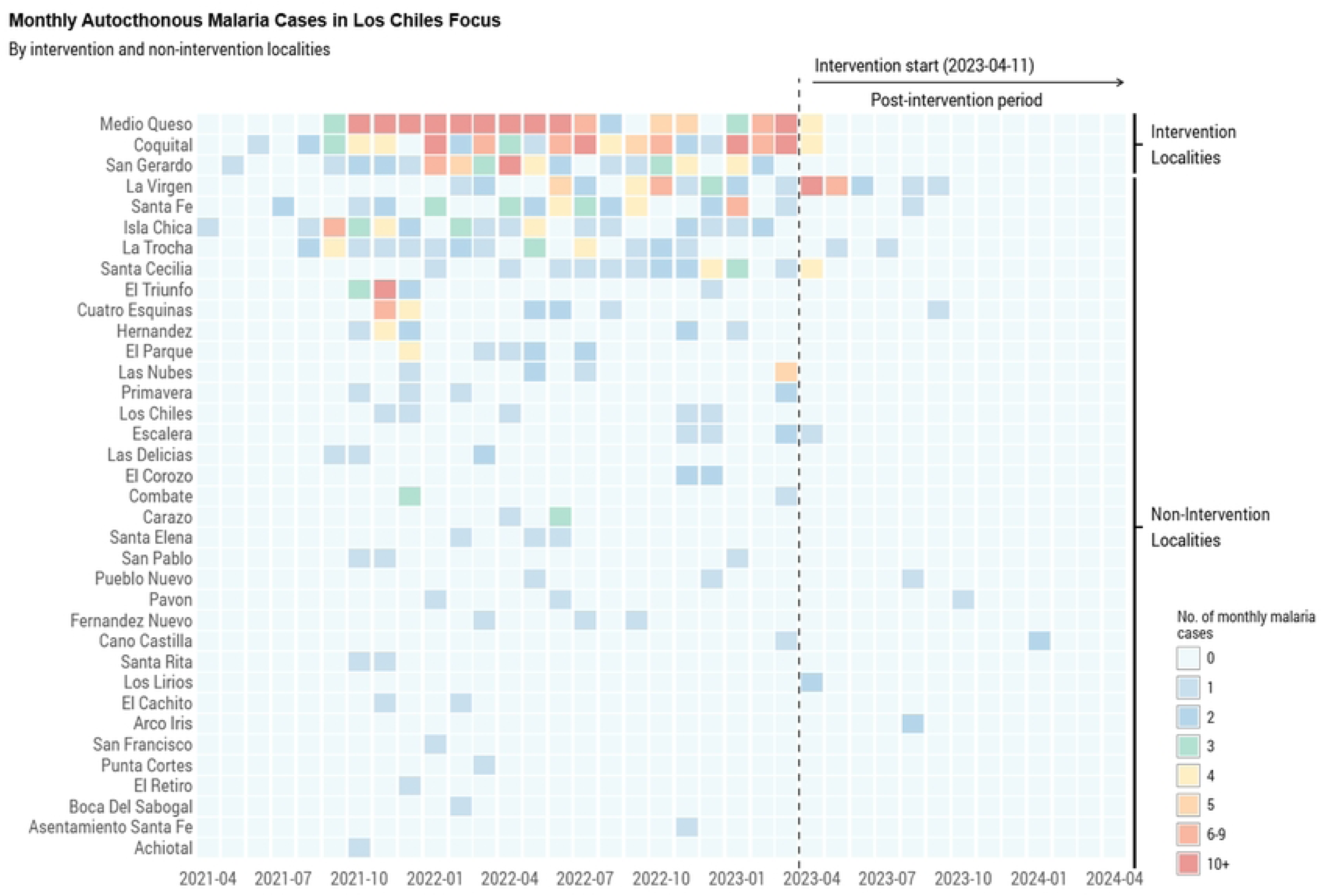
Heatmap of autochthonous malaria cases reported in the Los Chiles focus, Costa Rica, April 2021–April 2024.

Internal population mobility is a distinctive feature of this area. According to the Costa Rican Ministry of Health data, between April 2021 and March 2023, in 27.7% of confirmed cases, the probable place of infection within the Los Chiles focus did not correspond to the patient’s residence. Localities where the most people were infected outside their place of residence were Medio Queso (45 cases), Coquital (28), Santa Fe (14), San Gerardo (13), La Trocha (10), and La Virgen (10).

Considering the epidemiological situation, the Costa Rican Ministry of Health implemented the MDA strategy from April to June 2023 in the localities of Medio Queso, Coquital, and San Gerardo, as well as in two pineapple farms located near Medio Queso and Coquital. The persistent concentration of *P. falciparum* cases and the population characteristics supported the selection of MDA as a strategy with high potential to interrupt transmission in these localities [2].

The intervention involved the supervised administration of two cycles of chloroquine (25 mg/kg per cycle) to the eligible population, including people over one year of age without specific contraindications. Local malaria infections remain sensitive to chloroquine, which is the first-line treatment for malaria in Costa Rica, affecting all species (*P. falciparum*, *P. vivax*, and *P. malariae*). Each MDA cycle was administered over three consecutive days, with a seven-week interval between cycles. Before treatment, a door-to-door community engagement campaign was conducted to explain the objectives of the MDA, register eligible individuals, and coordinate the logistics of drug administration.

Prior to the implementation of the MDA, Costa Rica had taken steps to strengthen the capacity of its health system, particularly in high-priority areas such as the Los Chiles focus (see *Table 1*). Since 2019, the country has participated in the Regional Malaria Elimination Initiative (RMEI) [15], which aims to accelerate progress towards malaria elimination in Mesoamerica and the Dominican Republic. The initiative is funded by the Gates Foundation, the Carlos Slim Foundation, the Global Fund, and the nine participating countries, and is administered by the Inter-American Development Bank, in technical collaboration with the Pan American Health Organization.

**Table 1.**
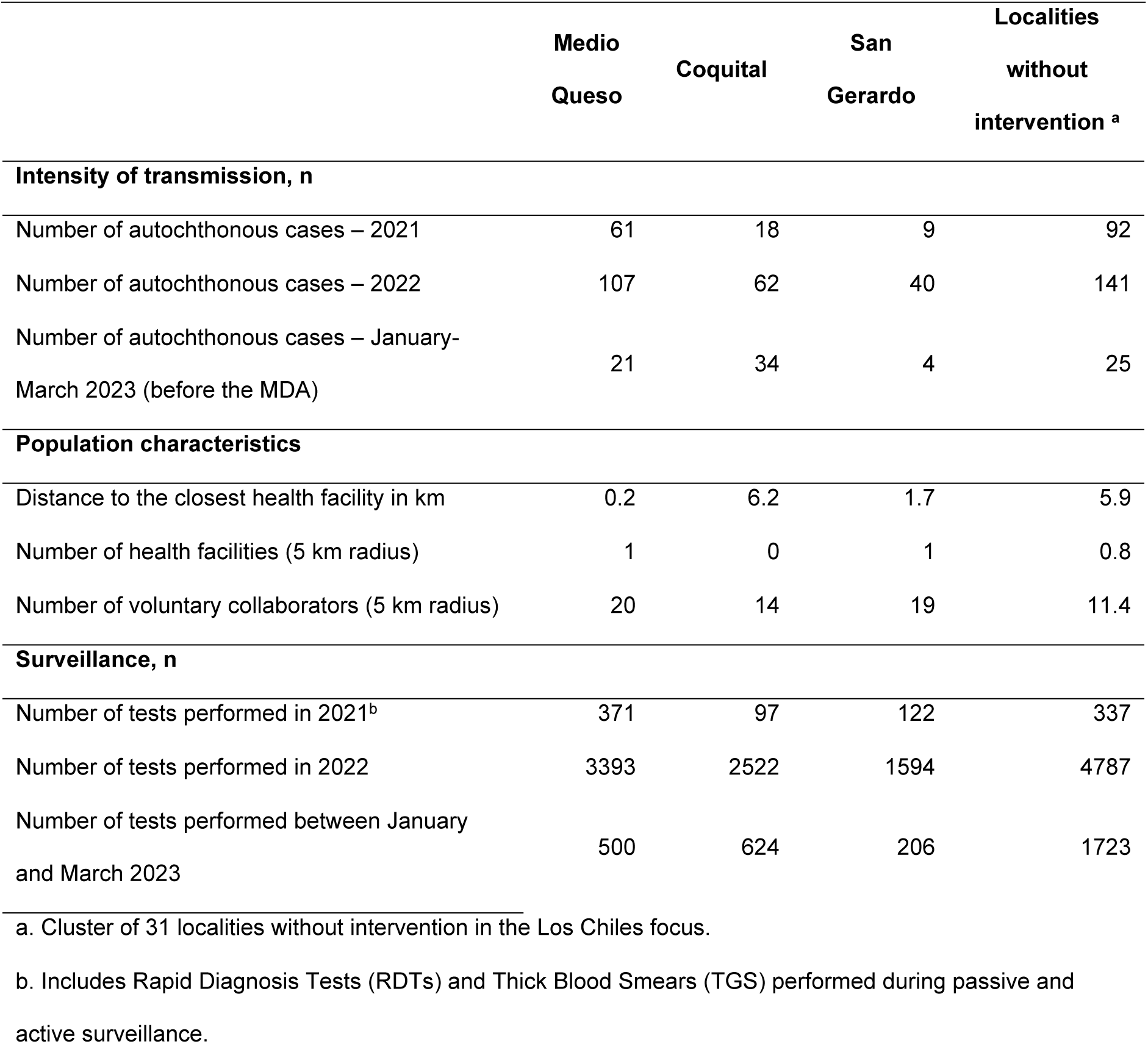
Characteristics of the localities in the Los Chiles Focus before Intervention.

|  | Medio<br>Queso | Coquital | San<br>Gerardo | Localities<br>without<br>intervention <sup>a</sup> |
| --- | --- | --- | --- | --- |
| **Intensity of transmission, n** |  |  |  |  |
| Number of autochthonous cases – 2021 | 61 | 18 | 9 | 92 |
| Number of autochthonous cases – 2022 | 107 | 62 | 40 | 141 |
| Number of autochthonous cases – January-March 2023 (before the MDA) | 21 | 34 | 4 | 25 |
| **Population characteristics** |  |  |  |  |
| Distance to the closest health facility in km | 0.2 | 6.2 | 1.7 | 5.9 |
| Number of health facilities (5 km radius) | 1 | 0 | 1 | 0.8 |
| Number of voluntary collaborators (5 km radius) | 20 | 14 | 19 | 11.4 |
| **Surveillance, n** |  |  |  |  |
| Number of tests performed in 2021<sup>b</sup> | 371 | 97 | 122 | 337 |
| Number of tests performed in 2022 | 3393 | 2522 | 1594 | 4787 |
| Number of tests performed between January<br>and March 2023 | 500 | 624 | 206 | 1723 |

a. Cluster of 31 localities without intervention in the Los Chiles focus.
b. Includes Rapid Diagnosis Tests (RDTs) and Thick Blood Smears (TGS) performed during passive and active surveillance.

As part of this initiative, Costa Rica implemented the Detection, Diagnosis, Treatment, Investigation and Response (DTI-R) strategy nationwide, supported by malaria stratification and foci management efforts. This included the integration of health services with vector control personnel in the processes of case detection, diagnosis, and treatment, as well as the consolidation of passive surveillance. The initiative also promoted the introduction and expansion of rapid diagnostic tests to enhance diagnostic capacity and improve the timeliness of case detection. The diagnostic network was expanded through the incorporation and consolidation of voluntary collaborator networks. Additionally, focus management teams were established to coordinate and implement the DTI-R strategy, tailored to the specific epidemiological conditions of the Los Chiles focus [16].

In this context, our study has two objectives. First, to evaluate the impact of MDA on the incidence of autochthonous malaria cases in the targeted localities, compared to what would have occurred in the absence of the MDA. Second, to provide a detailed description of the MDA intervention implemented in the selected localities within the Los Chiles focus.

## Methods

### MDA Intervention

The MDA in the Los Chiles focus was implemented between April 11 and June 9, 2023, by the Huetar Norte Regional Health Direction, in collaboration with the Los Chiles Health Area of the Costa Rican Ministry of Health, with support from other regional directorates. The intervention consisted of two phases: a community engagement campaign phase followed by a drug administration phase, delivered in two cycles. It targeted the entire eligible population in the localities of Medio Queso, Coquital, and San Gerardo, as well as two nearby farms.

Eligibility criteria included being at least one year of age, having no history of hypersensitivity or allergy to chloroquine, seizures, psoriasis, or purpura, and not having received any treatment with chloroquine or hydroxychloroquine within the four weeks before the intervention. The drug was administered in a total dose of 25 mg/kg of chloroquine (CQ) in 150 mg tablets distributed over three consecutive days per cycle. Dosages were adjusted by age group, following national guidelines [16] (see Table 2).

**Table 2.** Full Treatment Dose per Cycle.

| Intervention (25mg/kg) |  |  |  |
| --- | --- | --- | --- |
| Age group<br>(years) | kg | Tablets CQ | mg CQ |
| 1 – 2 | 12 | 2 | 300 |
| 3 – 6 | 18 | 3 | 450 |
| 7 – 11 | 30 | 5 | 750 |
| 12 – 14 | 54 | 9 | 1350 |
| 15+ | 60 | 10 | 1500 |

kg: Kilograms; CQ: Chloroquine; mg: Milligrams

To maximize coverage, the MDA operational strategy was designed considering internal mobility and daily routines within the target localities. The community engagement campaign served as an opportunity to test field operations and gain deeper insights into population movements. These findings informed the assignment of geographic sectors to each team, the definition of work schedules and visit hours, and the establishment of communication channels among field coordinators to help reach individuals likely to move within and between locations during the drug administration period.

The community engagement campaign was conducted from April 11 to 13, 2023, spanning one day per location. The campaign employed a door-to-door approach, visiting multiple locations, including homes, businesses, farms, schools, and other sites. Its purpose was to communicate the objectives of the MDA, distribute informational materials, register eligible individuals, and refine the logistics for the drug administration cycles. The core message emphasized the importance of treating the entire population, including those who appeared healthy, rather than focusing solely on symptomatic individuals. This approach aimed to achieve community understanding and acceptance, thereby minimizing resistance to treatment.

A week later, the first MDA cycle was launched, taking place from April 17 to 21, 2023. The second cycle was conducted seven weeks later, from June 5 to 9, following the same operational approach (see *Fig 4*). The MDA was implemented simultaneously in the three target localities by two-person teams assigned to specific geographic sectors. Drug administration was conducted over five days (three days for supervised treatment and two additional days to catch up with anyone missing treatment doses) with efforts made to operate outside regular working hours to maximize coverage.

**Fig 4.**
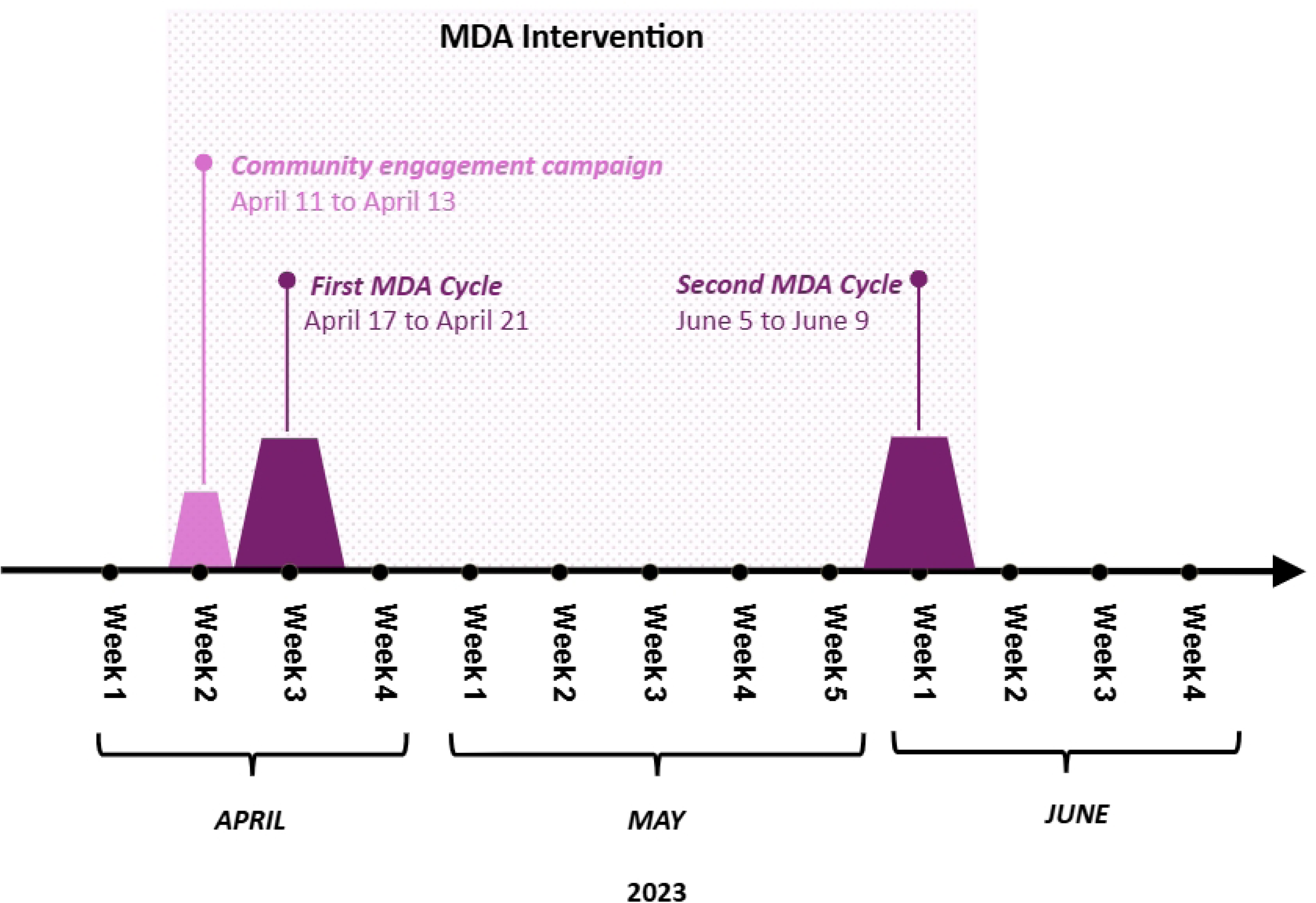
Intervention timeline.

After treatment, each individual was given a receipt to facilitate monitoring and tracking between cycles. On the two farms included in the intervention, the logistics differed: medication was administered on-site before the start of the workday to ensure that most workers received treatment.

Monitoring for adverse effects was conducted between cycles and following the second cycle. Any individuals who reported side effects were referred to local health centers for appropriate care. Volunteer collaborators supported these monitoring efforts within their communities; senior managers were present during kick-off and early implementation, including the Costa Rican Minister of Health and local leaders.

### Identification Strategy and Methods to Estimate the Causal Effect of MDA

We applied a target trial approach [17] to estimate the overall average effect of the MDA strategy on new autochthonous malaria cases in the three localities of Costa Rica. We defined the overall effect of MDA as the aggregate impact of the intervention on the outcome of interest across the entire target population [18]. This impact measure accounts for programmatic coverage, adherence to the antimalarial regimen, the direct pharmacological efficacy of the drugs administered, and the magnitude of any indirect effects resulting from the reduction in community transmission, all considered during the defined follow-up period.

#### Value and Importance of the Target Trial Approach

The target trial approach provides a robust conceptual and analytical framework for improving the estimation of causal effects from observational data, both in clinical settings [19,20] and in community-level or policy implementation contexts [21,22]. Its primary value lies in guiding observational research by explicitly specifying the protocol for a hypothetical randomized controlled trial (RCT), or the target trial, that would be designed to answer the causal question of interest if it was feasible to conduct it.

The process of defining the target trial requires rigorous clarification of all the study design components, including eligibility criteria, treatment strategies (intervention and comparator), the start of follow-up (time zero), outcome definitions, and the statistical analysis plan. Once the target trial protocol has been specified, the next step is to emulate it as closely as possible using available observational data.

The importance of this approach is multifaceted. First, by attempting to replicate the elements of an RCT, target trial emulation helps mitigate or make evident biases common in traditional observational studies, such as immortality bias, time-dependent selection bias, and indication confounding. Second, it promotes greater transparency in study design and assumptions, facilitating critical appraisal of results and reproducibility. Finally, by enforcing a clear definition of the causal question and the estimate of interest, it strengthens the validity of results from non-experimental data, which is crucial in settings where RCTs are not ethical, feasible, or generalizable to real-world settings.

#### Data sources

We obtained morbidity data from the Costa Rican National Malaria Surveillance System, which operates following national protocols [16]. This system compiles individual case data from malaria notification and investigation forms. The data include demographic information (sex, age, place of residence), clinical care details, diagnostic information (date of symptom onset, date of diagnosis, *Plasmodium* species, diagnostic method, place of diagnosis, probable place of infection), treatment received, and follow-up information. For our analysis, we used data on autochthonous cases reported between January 2021 and April 2024. We analyzed data by locality using 30-day time intervals for both the pre- and post-intervention periods.

Additionally, we used data on the number of malaria diagnostic tests conducted (including thick blood smears and rapid diagnostic tests) consolidated by the Ministry of Health. This dataset includes information on the date of testing, place of residence, health facility, diagnostic method, and case detection approach (passive or active). It encompasses tests conducted by the Costa Rican Social Security Fund, the country’s sole public healthcare provider, as well as those performed by vector control teams and community volunteers under the Ministry of Health.

We obtained detailed information on the MDA intervention (such as the schedule, components, target population, coverage, and adherence data) from data collected directly by the Costa Rican Ministry of Health. Before the intervention, local Ministry of Health teams conducted a census of the population in each locality to identify all eligible individuals. Subsequently, healthcare personnel and MDA teams documented individually for each cycle the doses received, any reported symptoms, and potential adverse effects using paper-based monitoring forms.

Finally, we consolidated data from various sources for each cycle, taking into account the movement of individuals within and between cycles and localities. This information was used to determine population coverage and adherence to the MDA intervention.

#### Target Trial Design and Emulation Using Observational Data

We describe the protocol of the target trial below. Table 3 presents a side-by-side comparison of the components of the hypothetical target trial protocol and our observational data-based emulation.

**Table 3.** Comparison between Target Trial and Emulation using Observational Data.

| **Protocol**<br>**Component** | **Target Trial:**<br>**Cluster-randomized clinical trial** | **Emulation of the Target Trial**<br>**using Observational Data** |
| --- | --- | --- |
| **Research**<br>**Question** | What is the overall average treatment effect on the treated (ATT), estimated under an intention-to-treat (ITT) framework, of a mass drug administration (MDA) strategy with antimalarials on the incidence of autochthonous malaria at 1, 3, 6, and 12 months after implementation, compared to no MDA implementation, in localities within the Los Chiles focus of Costa Rica? |  |
| **Eligibility** | **At the cluster level:** Localities within the Los Chiles malaria focus had to meet three conditions for inclusion: 1) report at least one autochthonous malaria case in the six months prior to randomization; 2) be located at least 5 km from any other selected locality to minimize interference; and, 3) have no overlap with other selected localities in terms of the case's residence or probable place of infection.<br>**At the individual level:** Eligible individuals were at least 1 year of age, and no history of hypersensitivity or allergy to chloroquine, seizures, psoriasis, or purpura, as well as no treatment with chloroquine or hydroxychloroquine within the 4 weeks prior to the MDA. | **At the cluster level:** For inclusion, eligible localities within the Los Chiles malaria focus were required to have reported at least one autochthonous malaria case in the six months prior to the intervention start date. The intervention group comprised three localities, San Gerardo, Medio Queso, and Coquital, where the MDA strategy was launched on April 11, 2023. Potential control localities had to meet the same initial eligibility criteria, be located at least 5 km from any intervention locality, and have no overlap with intervention sites of the case's residence or probable place of infection in the two previous years.<br>**At the individual level:** The same eligibility criteria as the target trial. |
| **Therapeutical interventions** | The intervention consisted of three sequential phases:<br>• Community engagement campaign | Same as target trial. |
|  | • First cycle of chloroquine administration (25 mg/kg), delivered as 150 mg tablets over three consecutive days, initiated one week after the engagement campaign<br>• Second cycle of chloroquine administration (25 mg/kg), also delivered over three consecutive days, conducted two months after the first cycle |  |
| **Treatment assignment** | Cluster-level random assignment of localities to either the intervention or control group. | Treatment assignment will be randomized, conditional on pre-treatment trends observed over a 24-month period, and adjusted for time-dependent confounding variables, including the number of imported cases, the number of thick blood smears performed, and the number of rapid diagnostic tests conducted, either by passive or active case detection.<br>Variables such as gender distribution, total population, educational attainment, and occupation are assumed to remain stable over time and are therefore excluded from adjustment, as they are |
|  |  | not expected to change meaningfully during the study period. |
| **Follow-up** | Time zero will be defined as the point of randomization.<br>All locations will be followed for 12 months. | Time zero will be defined as the start of the intervention for both the treatment and control groups.<br>Follow-up duration will mirror that of the target trial. |
| **Results** | Autochthonous malaria cases according to the place of infection and the date of symptoms onset. | Same as target trial. |
| **Causal contrasts** | Average overall treatment effect in those treated under intention to treat. | This study estimates the observational analogue of the average treatment effect on the treated (ATT) under an intention-to-treat (ITT) framework. It is assumed that the treated localities were those designated to receive the intervention, regardless of adherence at either the locality or individual level. |
| **Statistical Analyses** | To estimate the average treatment effect on the treated (ATT), both difference-in-differences (DiD) and the ratio of rate ratios (RRR) will be calculated, accounting for the clustered structure of the data. | The intention-to-treat ATT will be estimated using generalized synthetic control methods (GSCM). This approach generates weighted synthetic controls that serve as counterfactuals, representing the outcomes that would have been observed in the intervention |
|  | In a well-conducted and adequately powered cluster-randomized trial, the ATT is expected to approximate the average treatment effect (ATE). Therefore, simpler estimators, such as the crude rate ratio (RR) or the crude risk difference (RD), may also be used to estimate overall effects.<br>**Subgroup analyses:** Time-specific and location-specific effects will be estimating, allowing for the assessment of potential temporal and between-unit heterogeneity.<br>**Sensitivity analyses:** Given the risk of contamination (interference) effects, sensitivity analyses will explore the influence of geographic proximity and population overlap between treated and control localities. | group in the absence of the intervention. Model parameters will be selected based on pre-treatment model fit, ensuring optimal balance between intervention and control groups. 95% confidence intervals will be constructed using stratified bootstrapping.<br>**Subgroup analysis:** Same as target trial.<br>**Sensitivity analysis:** Analyses based on geographic proximity will include comparisons using control localities located within 5 km of intervention localities, as well as controls located outside the Los Chiles focus. In addition, potential contamination effects due to population overlap will be explored separately. This will be assessed through shared malaria case reporting patterns between intervention and control localities, based on patient residence or probable place of infection. |
|  |  | Proximity-based analyses are expected to reveal potential contamination effects. If such effects are present, comparisons using control locations in close proximity to intervention localities may be biased toward the null, reflecting the influence of interference between localities. In contrast, analyses using control localities outside the Los Chiles focus are hypothesized to be less affected by contamination, but they may provide a less comparable counterfactual due to potential differences in baseline characteristics, such as health system capacity.<br>Other analyses include implementing different estimators. |

The target trial protocol specifies clear eligibility criteria. At the cluster level, localities within the Los Chiles malaria focus had to meet three conditions for inclusion: report at least one autochthonous malaria case in the six months before randomization; be located at least 5 km from any other selected locality to minimize interference; and, have no overlap with other selected localities in terms of the case’s residence or probable place of infection. At the individual level, eligible participants comprised those who met the inclusion criteria for the MDA intervention.

In terms of treatment strategies, communities assigned to the control group would continue to receive the standard interventions already in place, including the DTI-R strategy, malaria testing with thick blood smears and rapid diagnostic tests, support from a network of community volunteers, and coordination by foci management teams. In contrast, the communities in the intervention group would receive the MDA intervention (including the engagement campaign and drug administration) in addition to the standard control group strategies.

Treatment assignment in the target trial would be conducted using cluster randomization, distributing eligible localities between the intervention and control groups. Follow-up in all participating localities would continue for 12 months, starting from time zero, defined as the time of randomization. The primary outcome of the study would be the number of autochthonous malaria cases, identified by locality of infection and date of symptom onset, assessed at 1, 3, 6, and 12 months following the start of the first MDA cycle.

The causal estimand of interest is the average treatment effect among those treated (ATT) under an intention-to-treat (ITT) approach at the locality level. This estimand compares the outcomes observed in localities assigned to the MDA intervention with the expected outcomes under non-implementation, hence, capturing the overall effect of the intervention at the locality level.

For statistical analysis, the primary approach involves estimating the ATT using difference-in-differences and ratio of rate ratios (RR) methods, accounting for clustering by locality. Additionally, subgroup analyses were planned to explore potential heterogeneity in treatment effects over time and across different locations. Finally, sensitivity analyses would assess the potential impact of interference, considering geographic proximity and population overlap between localities in intervention and control groups.

#### Emulation of the Target Trial using Observational Data

To emulate the target trial using observational data, we applied the same eligibility criteria at the cluster level for the localities within the Los Chiles focus. We defined intervention localities (San Gerardo, Medio Queso, and Coquital) as those where the MDA strategy was initiated on April 11, 2023, and where at least one autochthonous malaria case had been reported in the six months preceding the intervention. Potential control localities had to meet the same malaria history criteria as the target trial: be located at least 5 km from any intervention site and have no overlap in malaria case reporting (based on place of residence or probable place of infection) with the intervention localities. At the individual level, we maintained the same eligibility criteria as those specified for the target trial protocol.

Regarding treatment strategies, the intervention implemented in the treated localities replicated the three-component structure of the target trial: a community engagement campaign, followed by two supervised drug administration cycles. The control localities continued with standard activities, without the MDA implementation.

Treatment assignment was assumed to follow conditional randomization. This assumption was supported by pretreatment trends in the primary outcome observed over a 24-month period and on a series of specific time-varying confounders; the number of imported malaria cases and the volume of thick blood smears and rapid diagnostic tests performed under both passive and active case detection strategies. Due to the limited follow-up period, we considered covariates such as gender distribution, total population, educational levels, and occupation as time-invariant and excluded them from adjustments.

We defined time zero as April 11, 2023, the start of the intervention, for both treatment and control groups. The follow-up period was extended for 12 months, analogous to the target trial. The primary outcome measure was also the same as the target trial: the autochthonous malaria case count, defined by the location of infection and the date of symptom onset. We sought to estimate the observational causal contrast analogue of the AT under an ITT approach, assuming that the treated localities were the three intervention localities, without considering adherence levels within localities or between individuals.

#### Statistical Analysis

Our primary analysis focused on estimating the ATT under ITT using generalized synthetic control methods (GSCM), since the parallel trends assumption, required for methods such as difference-in-differences, was not expected to hold. We implemented this method with the *gsynth* package [23] in R [24], which applies matrix completion techniques to impute potential missing outcomes (i.e., counterfactuals) for treated units [25]. In essence, this method simulates the untreated scenario for intervention localities using information from comparable control units. We selected the model hyperparameters through cross-validation using the predicted minimum mean squared errors as the selection metric. We calculated 95% confidence intervals for the estimated differences and ratios using 1000 stratified bootstrap iterations.

We conducted subgroup analyses to estimate specific effects at different time points and across different locations to investigate potential heterogeneity in effects, in line with the target trial. We also performed comprehensive sensitivity analyses. To address the risk of interference, we explored the impact of geographic proximity and population overlap by comparing outcomes using controls located within 5 km of treated units versus controls outside the Los Chiles focus. This aims to asses the potential bias toward the null due to interference by using nearby controls versus the reduced comparability when using more distant controls. Additionally, we performed sensitivity analyses using alternative estimators, such as two-way fixed effects difference-in-differences and conventional differences-in-differences using linear regression.

### Ethics Statement

This study was approved by the National Health Research Council of Costa Rica (Consejo Nacional de Investigación en Salud, CONIS), under agreement 10/2024 (letter No. CONIS-448-2024). Our research involved secondary analysis of identifiable data, accessed with appropriate institutional authorization and in compliance with national data protection laws. Strict measures were taken to ensure data confidentiality and secure storage. All necessary ethical safeguards were implemented, and the use of identifiable information was justified and approved by the ethics committee.

## Results

### MDA Coverage and Adherence

We found that of the 4,624 individuals registered across the three localities, 4,316 (93.3%) received at least one dose of chloroquine in at least one MDA cycle. Among the total population, 35.2% participated in both cycles, 42.1% only in the first cycle, 16.0% only in the second cycle, and 6.7% did not participate in either cycle. Coverage with at least one dose by cycle was 77.3% in the first cycle and 51.2% in the second cycle (see Table 4). Regarding adherence, 68.1% of the total registered individuals received a full treatment course in at least one cycle. Specifically, 20.4% completed treatment in both cycles, 37.6% only in the first cycle, 10.1% only in the second cycle, and 31.9% either did not participate or did not complete treatment in either cycle.

**Table 4.**
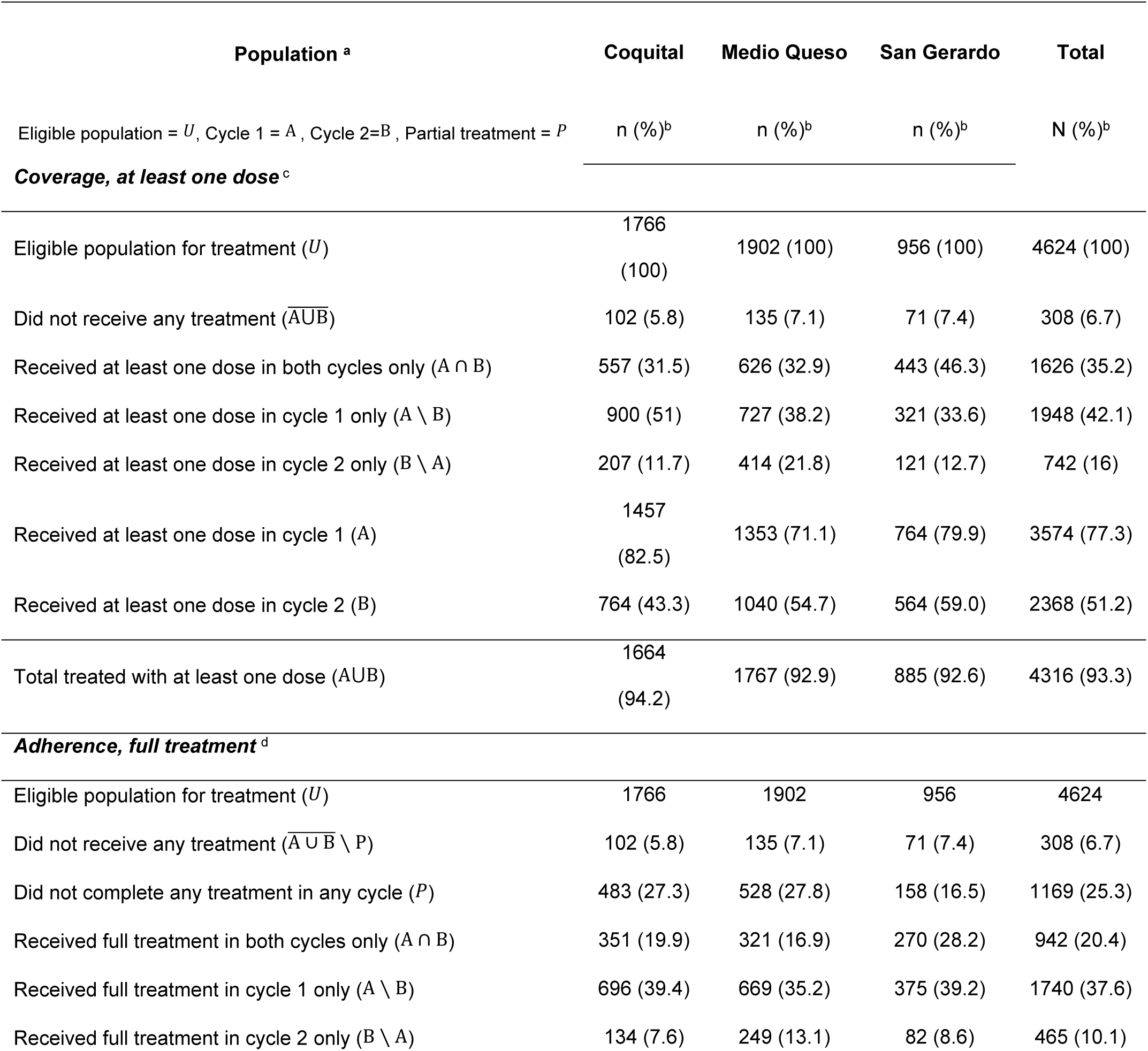
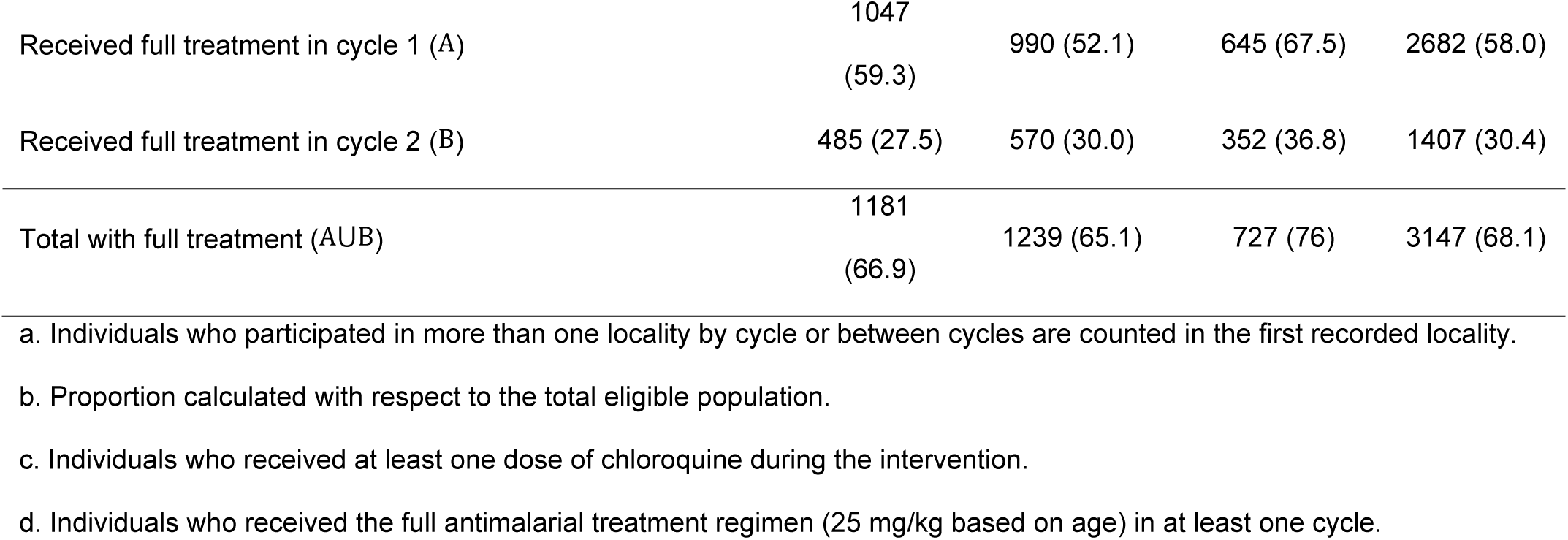
MDA Coverage and Adherence by Locality.

Across the three localities, MDA coverage with at least one dose ranged from 92% to 94% considering both cycles, with complete treatment in at least one cycle exceeding two-thirds of the population. Coquital achieved the highest coverage in the first cycle, with 82.5% individuals receiving at least one treatment dose; however, coverage in the second cycle declined to 43.3% (see Table 4). In contrast, San Gerardo had the highest proportion of individuals who received full treatment in at least one cycle (76.0%) and led in overall coverage and full treatment across both cycles. This was followed by Coquital (66.9%) and Medio Queso (65.1%) (see Table 4).

Internal mobility within and between localities was reflected in the coverage data: 61 individuals received treatment in different localities in the first cycle (e.g., the first dose in Coquital and subsequent doses in Medio Queso), six during the second cycle, and 81 individuals received treatment in different localities across the two cycles (see Table 5).

**Table 5.**
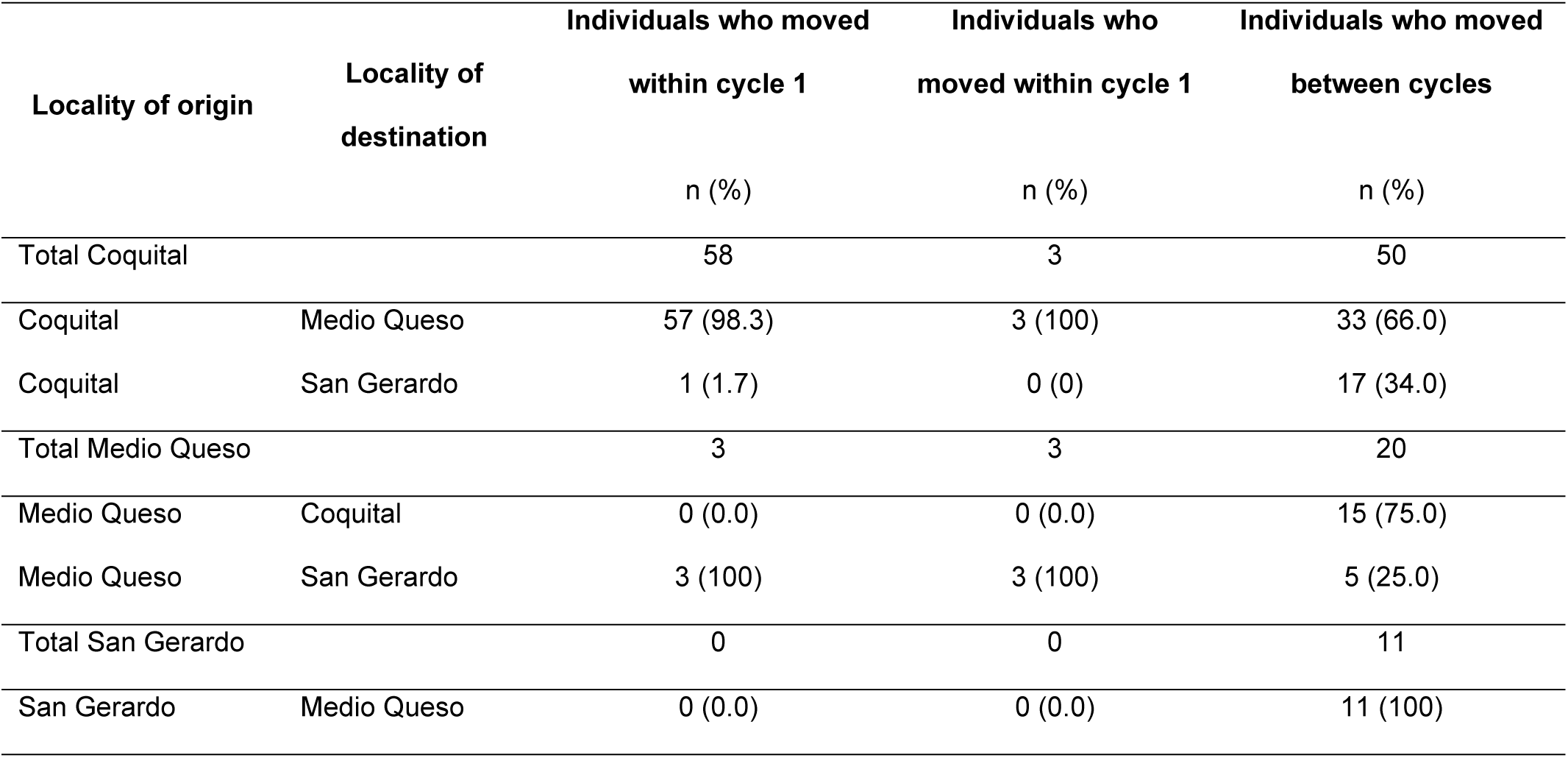
Population movement between localities during MDA cycles.

| Locality of origin | Locality of destination | Individuals who moved | Individuals who | Individuals who moved |
| --- | --- | --- | --- | --- |
|  |  | within cycle 1 | moved within cycle 1 | between cycles |
|  |  | n (%) | n (%) | n (%) |
| Total Coquital |  | 58 | 3 | 50 |
| Coquital | Medio Queso | 57 (98.3) | 3 (100) | 33 (66.0) |
| Coquital | San Gerardo | 1 (1.7) | 0 (0) | 17 (34.0) |
| Total Medio Queso |  | 3 | 3 | 20 |
| Medio Queso | Coquital | 0 (0.0) | 0 (0.0) | 15 (75.0) |
| Medio Queso | San Gerardo | 3 (100) | 3 (100) | 5 (25.0) |
| Total San Gerardo |  | 0 | 0 | 11 |
| San Gerardo | Medio Queso | 0 (0.0) | 0 (0.0) | 11 (100) |

### MDA Effectiveness

#### Overall Effect of MDA on Autochthonous Malaria Cases

Following the MDA intervention, the number of observed autochthonous malaria cases in treated locations dropped to zero in the first 30 days and remained at zero throughout the 12-month follow-up period. In contrast, the GSCM estimated that cases would have persisted, fluctuating between approximately 3 and 4 cases per 30-day block. Fig 5 shows the mean number of observed autochthonous malaria cases across all intervention localities, compared with the synthetic control estimates, both before and after MDA implementation.

**Fig 5.**
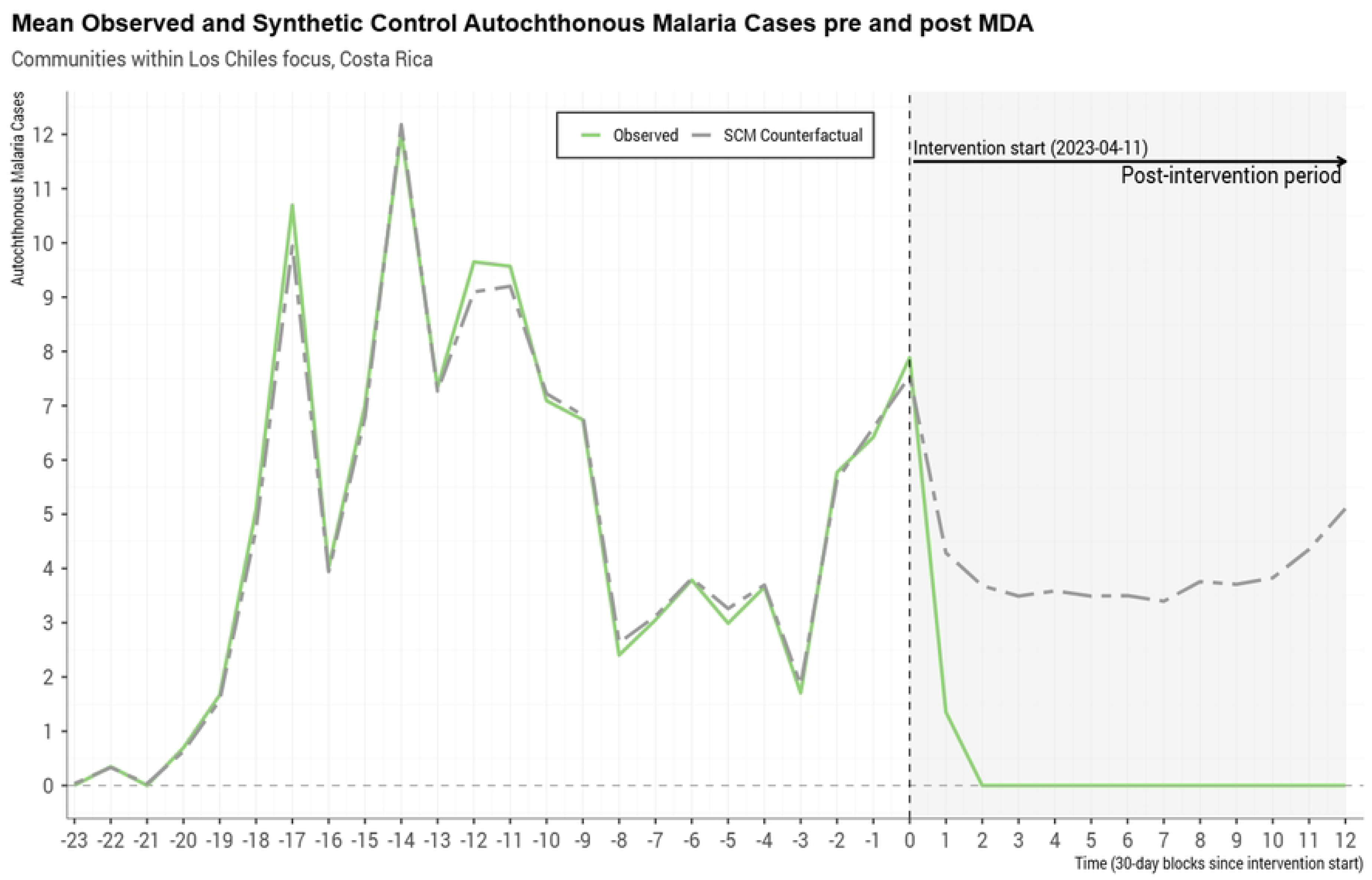
Average number of autochthonous malaria cases observed versus estimated using synthetic controls.

Over the entire post-intervention period, the MDA strategy was associated with a significant reduction in malaria cases. The overall rate ratio (RR) was 0.23 (95% CI: 0.13, 0.49), indicating a 77% reduction in the rate of new cases. The absolute difference showed an average reduction of −3.72 cases per period (95% CI: −7.2, −0.94) relative to the synthetic control. Table 6 summarizes the average treatment effect on the treated cases for all intervention localities.

**Table 6.** Average global effect of the intervention on the treated.

|  | t1 | t3 | t6 | t12 | Global post-intervention period |
| --- | --- | --- | --- | --- | --- |
| **Global** |  |  |  |  |  |
| RR [IC 95%] | 0.46 [0.2, 1.02] | 0.21 [0.12, 0.47] | 0.21 [0.12, 0.52] | 0.19 [0.11, 0.5] | **0.23 [0.13, 0.49]** |
| RD [IC 95%] | -2.74 [-6.63, 1.05] | -3.72 [-7.21, -1.12] | -3.68 [-7.16, -0.93] | -4.24 [-8.2, -1.02] | **-3.72 [-7.2, -0.94]** |
| **Coquital** |  |  |  |  |  |
| RR [IC 95%] | 0.46 [0.22, 0.49] | 0.22 [0.12, 0.32] | 0.22 [0.12, 0.33] | 0.22 [0.12, 0.37] | **0.24 [0.13, 0.33]** |
| RD [IC 95%] | -2.35 [-6.9, -2.07] | -3.6 [-7.55, -2.12] | -3.59 [-7.54, -2.04] | -3.64 [-7.54, -1.73] | **-3.49 [-7.48, -2.26]** |
| **Medio Queso** |  |  |  |  |  |
| RR [IC 95%] | 0.49 [0.43, 1.06] | 0.15 [0.12, 0.35] | 0.15 [0.13, 0.34] | 0.12 [0.07, 0.17] | **0.17 [0.15, 0.37]** |
| RD [IC 95%] | -4.17 [-5.21, 0.22] | -5.86 [-7.02, -1.89] | -5.79 [-6.99, -1.94] | -7.42 [-14.37, -5.06] | **-6 [-7.03, -2.46]** |
| **San Gerardo** |  |  |  |  |  |
| RR [IC 95%] | 0.37 [0.21, 0.78] | 0.37 [0.23, 0.5] | 0.38 [0.23, 0.58] | 0.38 [0.23, 0.52] | **0.37 [0.23, 0.56]** |
| RD [IC 95%] | -1.71 [-3.89, 1.06] | -1.71 [-3.29, -0.99] | -1.65 [-3.28, -0.73] | -1.64 [-3.27, -0.91] | **-1.69 [-3.28, -0.83]** |

RR: Rate ratio; RD: Rate difference

The effect over time indicates a sustained protective effect following the MDA intervention. At 1-month post-intervention (t_1_), the RR was 0.46 (95% CI: 0.2, 1.02), indicating a reduction that was not statistically significant. However, from 2 months onward, the reductions were statistically significant. At 3 months (t₃), the RR was 0.21 (95% CI: 0.12–0.47), with an absolute difference of –3.72 cases (95% CI: –7.21 to –1.12). At 6 months (t₆), the RR remained at 0.21 (95% CI: 0.12–0.52), with a difference of –3.68 cases (95% CI: –7.16 to –0.93). By 12 months (t₁₂), the RR further declined to 0.19 (95% CI: 0.11–0.50), corresponding to an absolute reduction of –4.24 cases (95% CI: –8.20 to –1.02).

#### Heterogeneity of MDA Effects Across Localities

Differences in MDA effects were observed across the three intervention locations, although all showed substantial reductions in malaria incidence (see Table 6, Fig 6 and 7).

**Fig 6.**
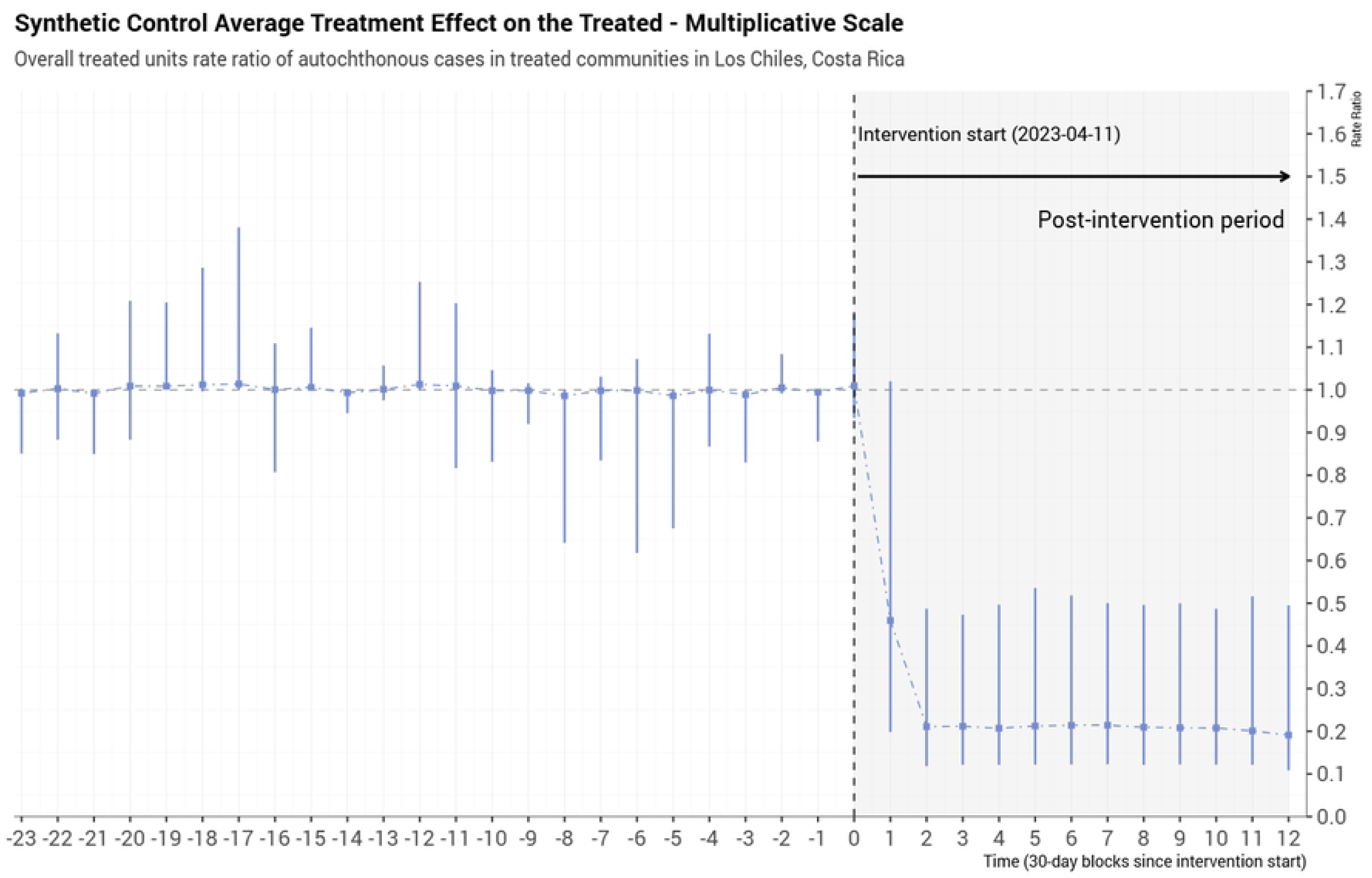
Overall treatment effect on the treated, estimated using synthetic controls.

**Fig 7.**
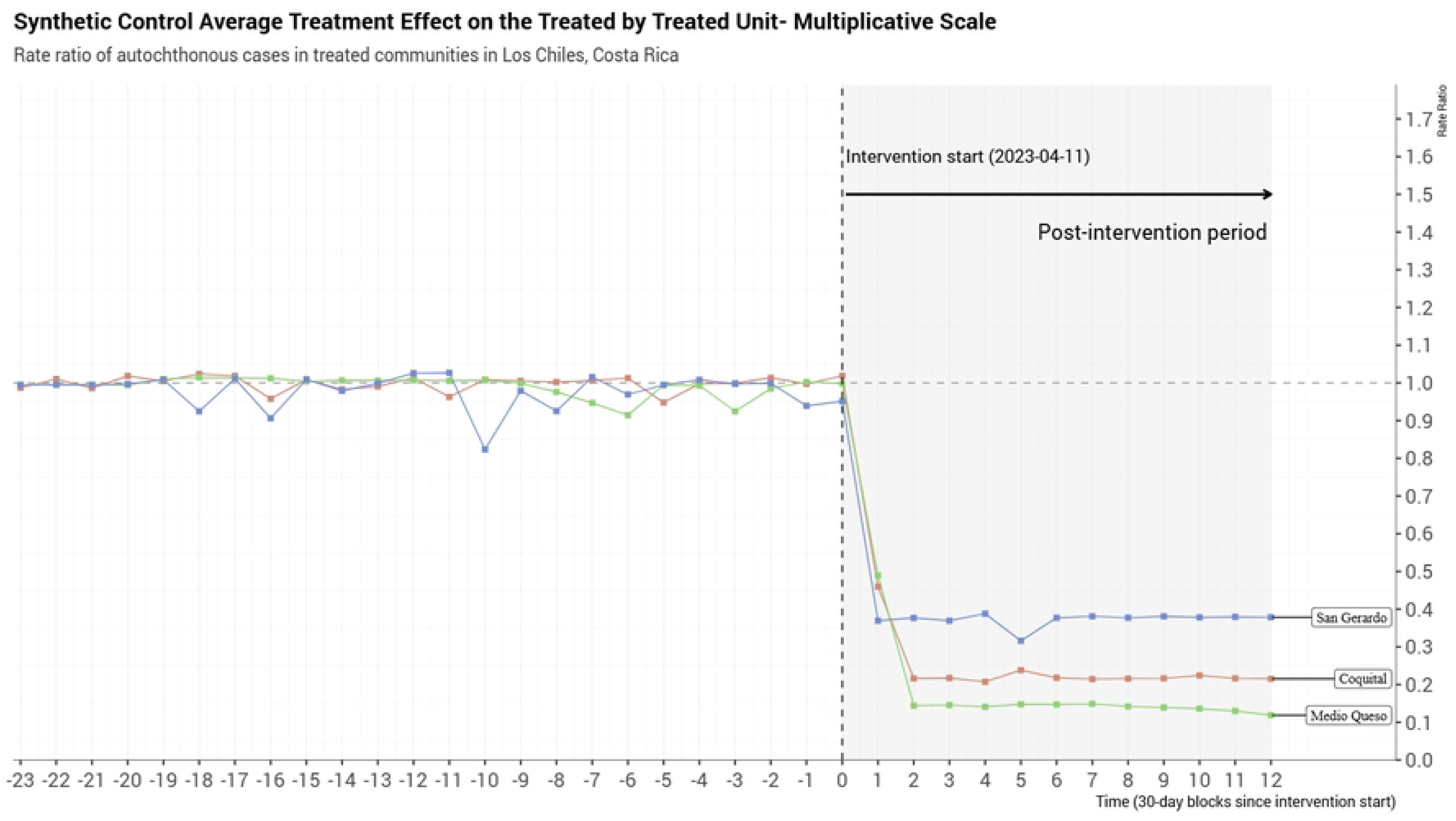
Treatment effect on the treated, estimated using synthetic controls, by locality.

In Coquital, the overall post-intervention RR was 0.24 (95% CI: 0.13–0.33), with an average reduction of –3.49 (95% CI: –7.48 to –2.26) cases per period. Statistically significant reductions were observed from t₁ (RR = 0.46; 95% CI: 0.22–0.49) through t₁₂ (RR = 0.22; 95% CI: 0.12–0.37).

In Medio Queso, the MDA intervention yielded the greatest relative reduction in incidence, with an overall RR of 0.17 (95% CI: 0.15–0.37) and an average difference of –6.00 (95% CI: –7.48 to –2.26) cases. The effect became statistically significant from t₂ onward (RR = 0.14; 95% CI: 0.12–0.34).

In San Gerardo, the overall post-intervention RR was 0.37 (95% CI: 0.23–0.56), corresponding to a difference of –1.69 (95% CI: –3.28 to –0.83) cases. Statistically significant reductions were observed from t₁ (RR = 0.37; 95% CI: 0.21–0.78) through t₁₂ (RR = 0.38; 95% CI: 0.23–0.52).

#### Sensitivity Analysis

We conducted a sensitivity analysis to assess the robustness of the estimated treatment effect under different control groups specifications used to construct the synthetic control counterfactual. Results are summarized in Table 7. In our primary analysis, control localities were selected within the Los Chiles focus, located more than 5 km from the treated localities and with no population overlap. This resulted in a post-intervention RR of 0.23 (95% CI: 0.13, 0.49). When restricting the control group to nearby or overlapping populations, the estimated effect was slightly attenuated (RR = 0.27, 95% CI: 0.19–0.44). Our findings are consistent with positive interference between treatment localities and the localities that are closest or with overlapping populations in the control group, which would bias the estimated treatment effect toward the null.

**Table 7.** Sensitivity Analysis by control group localities.

| Control donor pool group | RR [95% CI] |  | RD [95% CI] |  |
| --- | --- | --- | --- | --- |
| Los Chiles Focus: Localities < 5km from a treated locality or with population overlap |  | 0.27 [0.19, 0.44] |  | -3.01 [-4.42, -1.28] |
| Los Chiles Focus: Localities > 5km from a treated locality and without population overlap |  | 0.23 [0.13, 0.49] |  | -3.72 [-7.2, -0.94] |
| Los Chiles Focus: All localities |  | 0.24 [0.19, 0.47] |  | -3.56 [-4.87, -1.15] |
| Limon Province Foci: El Jardín and Limon |  | 0.13 [0.08, 0.2] |  | -7.16 [-11.51, -4.73] |
| RR: Rate Ratio; RD: Rate Difference; 95% CI: 95% Confidence interval |  |  |  |  |

Expanding the control pool to include all locations within the Los Chiles focus, regardless of proximity or overlap, yielded a similar estimate (RR=0.24, 95% CI: 0.19–00.47), again slightly attenuated compared to the primary analysis.

Finally, using an external an external control pool comprised of malaria foci from the Limón province, resulted in a more pronounced treatment effect. The post-intervention RR was 0.13 (95% CI: 0.08–00.2), and the absolute difference indicated an average reduction of 7.16 cases per period (95% CI: −11.51 to −4.73). While this suggests a larger effect, potentially less influenced by contamination, it may also reflect differences in baseline characteristics between Limón and Los Chiles that could affect comparability.

We also performed additional sensitivity analyses using alternative estimators. Results from these analyses were consistent with the primary findings, further supporting the robustness of the estimated treatment effect (see Table S1).

## Discussion

To our knowledge, this is the first MDA implementation study to apply an emulated target trial approach with a 12-month follow-up period. Our results demonstrate that the chloroquine-based MDA strategy implemented in the Los Chiles focus, Costa Rica, was highly effective, achieving a complete and sustained reduction in autochthonous malaria cases to zero throughout the 12-month follow-up. This outcome is particularly significant for malaria elimination efforts, both nationally and across the Central American region, as it indicates a sustained interruption of local transmission.

The main finding of our study, the complete interruption of *P. falciparum* malaria transmission for one year following two cycles of chloroquine-based MDA, stands out in the context of MDA research. Several systematic reviews [4,6,27] have documented that the MDA can significantly reduce malaria prevalence in the short term, especially in low-transmission settings, typically showing reductions in the three months following implementation. However, they found limited evidence on the durability of these effects beyond that period. In this context, the persistence of zero autochthonous cases for a full year in our study is a particularly encouraging result.

Several factors may have contributed to the sustained interruption of transmission. These include the initial low transmission intensity in the focus; prior efforts to strengthen local health system capacity, which enabled timely identification of autochthonous and imported cases; the high first-cycle coverage (77.3%); the potential continued efficacy of chloroquine in this population, where drug resistance may still be low; and a decline in malaria cases in neighboring areas of Nicaragua, where the national case count dropped by more than half between 2022 and 2023 [13].

The success of the MDA intervention is noteworthy given the operational challenges faced. While coverage in the first cycle was high, participation declined during the second cycle, anecdotally attributed to factors such as hesitancy due to adverse effects, population mobility, and shifts in the migrant labor population. The use of non-sugar-coated chloroquine tablets, with their bitter taste, may have also affected uptake and contributed to gastrointestinal side effects. Despite this reduced participation in the second cycle, transmission was still interrupted. This suggests that the intensity of the initial cycle may have been sufficient to markedly reduce the parasite reservoir in this low-transmission setting, even in the absence of full adherence in both cycles.

Moreover, the second cycle played a valuable role in expanding the intervention’s reach. It increased the proportion of individuals who received at least one dose by 16.0% and added a 10.1% increase in full treatment adherence.

A crucial operational innovation of this MDA strategy was its ability to provide treatment to mobile individuals, a known challenge for malaria elimination in Latin America. This flexibility was made possible using treatment receipts and effective coordination among drug administration teams across localities, enabling the identification and follow-up of individuals who moved between locations. Although the parasites in this region remain sensitive to chloroquine [28], the potential introduction of resistant strains, along with the need to monitor adverse effects, underscores the importance of implementing active pharmacovigilance and maintaining transparent communication about risks and benefits in future MDA campaigns.

The choice of chloroquine is relevant due to its potential efficacy in this specific epidemiological context and its very low cost, as well as its single daily dosage, a key factor in planning large-scale public health interventions in resource-limited settings. These findings provide valuable evidence on the feasibility of cost-effective MDA regimens tailored to particular epidemiological and operational conditions.

Our results also align with WHO and PAHO recommendations supporting MDA in elimination or pre-elimination settings, where a robust health system is essential [4,5]. Before the MDA, Costa Rica’s Ministry of Health and CCSS had significantly strengthened health system capacity in the Los Chiles region. This groundwork likely played an essential role in achieving the observed results. Sensitivity analyses indirectly support this notion by suggesting that the measured impact may reflect both the effect of MDA and the benefits of implementing it in the context of an effective health system. These findings underscore that MDA is most effective when integrated into a broader package of interventions and delivered within a well-supported, strengthened health system.

The strength of conclusions reinforced by the use of target trial emulation with synthetic controls [29]. This approach enables more rigorous causal inference from observational data by explicitly specifying the ideal trial to be conducted and emulating it using real-world data, mitigating common biases [30,31]. The consistency of our results across multiple sensitivity analyses, including variations in control group composition and the application of alternative statistical estimators, further supports the validity of our conclusions. Interestingly, estimates were slightly attenuated when using geographically proximate or overlapping populations in control localities, indicating positive contamination effects towards neighboring areas, which is a plausible phenomenon in community-based interventions of this nature.

Nonetheless, our study also has several limitations. First, because the intervention was not randomized, we cannot entirely exclude the possibility of unmeasured confounding factors that may have influenced the results. However, the robustness of the findings across the primary analysis and the multiple sensitivity analyses provides reassurance regarding the validity of the results. In terms of data quality, although studies relying on routine administrative data may face variability, the malaria incidence data used in this study were obtained from the official national surveillance system. Information on intervention coverage and adherence was collected directly by the Ministry of Health, using structured and standardized procedures. This suggests that the data were systematically compiled. Finally, our population estimates were derived from the MDA census and may not fully capture mobile populations, such as migrants, farmers, and others.

Generalizability of our findings requires caution. The use of synthetic control methods allows for estimating the ATT under an ITT framework at the cluster level. By design, the ATT reflects the effect of the MDA specifically in the three treated localities in the context of their unique epidemiological and operational conditions. Therefore, direct extrapolation of these findings to malaria foci with different characteristics (such as higher transmission intensity, existing *P. vivax* transmission, or less developed health systems) should be done with caution. Finally, although the interruption of *P. falciparum* transmission in Los Chiles is encouraging, its long-term sustainability will depend on the health system’s ability to promptly detect, diagnose, and treat any imported cases. Maintaining this capacity is crucial to preventing the re-establishment of malaria transmission.

## Conclusion

We demonstrate that the chloroquine-based MDA strategy implemented in three localities within the Los Chiles focus supported the interruption of autochthonous malaria transmission over a 12-month period. This experience in Costa Rica highlights the potential of MDA, when carefully planned and executed within a strengthened health system and in a low-transmission setting, to serve as a powerful tool to accelerate malaria elimination

Our findings have important implications for elimination strategies in Costa Rica and may provide a model for interventions in other priority areas with similar epidemiological conditions across Central America. Given the regional dynamics of malaria transmission, sustained cross-border collaboration will be essential to maintain these achievements.

To further consolidate these results, future research should prioritize evaluations on the cost-effectiveness of MDA, assessing its long-term impact beyond 12 months, and the adaptation of this model to other transmission settings, considering local epidemiological contexts and community acceptability. Continued operational research will be key to refining and optimizing malaria elimination strategies in Costa Rica and the broader region.

## Data Availability

Limited anonymized datasets supporting the findings of this study are available via the Open Science Framework (OSF) at: https://osf.io/tk7s3/?view_only=25021eb260754252826a2bdb620b39c0. These datasets include aggregated and de-identified information derived from malaria surveillance and intervention monitoring. The full, individual-level datasets are owned by Costa Rica's Ministry of Health and are not publicly available due to data protection regulations. Interested researchers may request access to these data directly from Costa Rica's Ministry of Health, subject to institutional and ethical approvals.

## Author contributions

MDA protocol development: LV, IVR, HHB, JGL, RMR, YCR; Conceptualization: ATM, RMR, MRR, IVR, YCU, HHB, JGL, AO, SE, DR; Data curation: AO, SE; Formal Analysis: AO, SE; Investigation: AO, SE; Methodology: AO, SE, DR; Project administration: ATM, MRR, YCU; Resources: ATM, RMR, MRR, IVR, YCU, HHB, JGL; MDA Field work: HHB, JGL, IVR, RMR, MRR, YCR, LV; Supervision: SE, DR; Validation: ATM, RMR, MRR, IVR, YCU, HHB, JGL, AO, SE, DR; Visualization: AO, SE; Writing – original draft: AO, SE, DR; Writing – review & editing: ATM, RMR, MRR, IVR, YCU, HHB, JGL, AO, SE, LV, DR.

